# Lysosomal Enzyme activity: Establishing Reference Intervals from Patient Data

**DOI:** 10.1101/2024.04.11.24303575

**Authors:** Dylan Mordaunt, Samantha Stark, Michael Fietz, Janice Fletcher, Michael Metz

## Abstract

**Objectives:** To develop reference intervals for leukocyte lysosomal enzymology.

**Design:** Standards-based establishment of reference intervals by analysis of 7851 for 17 assays of 15 lysosomal enzymes in leukocyte and plasma (January 1985 through September 2013).

**Methods:** Reference intervals were calculated using MedCalc for Windows version 12.5 (MedCalc Software, Ostend, Belgium), according to the Clinical and Laboratory Standards Institute C-28A protocol (CSLI-C28A).

**Results and Conclusion:** Evidence based reference intervals are presented in tabular form for seventeen assays of fifteen lysosomal enzymes.

## 1. Introduction

Lysosomal storage disorders (LSDs) are a group of genetic diseases involving accumulation of lipid and glycan molecules in the lysosome. All but two LSDs are inherited in an autosomal recessive manner, with alpha-galactosidase and iduronate-2-sulfatase deficiencies, known as Fabry and Hunter disease respectively, being X-linked. Although LSDs are individually rare disorders, collectively they represent an important differential diagnosis in the work-up of causally treatable genetic disease. Some of these disorders occur more commonly in certain populations, such as the prototypic neuronopathic LSD, beta-hexosaminidase A deficiency (Tay-Sachs disease or GM2 gangliosidosis). This disorder occurs more commonly in Ashkenazi Jews, Amish communities, and those of French-Acadian descent such as Quebec French-Canadians and the Cajun population of Louisiana.

Although making these diagnoses is an important milestone for any of these affected individuals and families, including the ability to inform reproductive and genetic counseling, the primary importance of detecting these disorders is in the availability of causal treatments for some of these disorders [1,2]. Treatments for these disorders can be conceptually categorized into substrate reducing [3] or enzyme-replacing therapies [4,5]. Predominantly forms of enzyme replacement are in use, such as allogeneic haematogenous stem cell transplant (HSCT) or infusions of recombinant humanized enzyme replacement therapies (rhERT) [4,5]. Therapies in development include gene therapies and small molecule therapies such as chaperone molecules [6,7].

For over 30 years, our laboratory has performed a panel of lysosomal enzyme assays (white cell enzymes, WCE’s) on leukocytes and plasma to diagnostically screen for a number of neuronopathic LSDs. Whilst a number of factors can affect measured enzyme activities in these patients, WCE activities outside the reference intervals can be verified by measurement of activity in cultured skin fibroblasts and increasingly with measurement of plasma/urine metabolites or gene analysis. Therefore, the WCE panel has enabled early diagnosis of these neuropathic LSDs, before the stereotypical symptoms and signs (which usually present late) occur [8,9]. Despite the revolution in gene-based diagnostics, enzymology continues to be important in diagnosis [10]. Response to treatments can also be verified by monitoring the specific white cell or plasma enzyme, substrate, metabolite or some secondary biomarkers such as macrophage activation (assessed via plasma chitotriosidase activity).

Seventeen lysosomal enzymes (fourteen in leukocytes and three in plasma) are currently measured in our laboratory, by fluorometry, spectrometry or radionuclide analysis. Our current reference intervals are unreferenced. We simply do not know how they were derived. Thus, our aim was to improve the quality of our laboratory’s performance and develop valid reference intervals.

The Stockholm Hierarchy is a professional consensus created in 1999 to define the preferred approaches to defining analytical quality [11]. Application of the Stockholm Hierarchy to reference intervals suggests that reference intervals derived from patient data are less desirable than reference intervals derived from clinical outcome studies [12]. However useful clinical outcome studies are not often available. In a priori reference interval studies, a reference population is defined first and then the analyte of interest is measured and reference intervals are described. Commonly a healthy population is chosen as the reference population, but a population with characteristics similar to the population affected by the disease but free of the disease can provide more useful reference intervals. [12] A posteriori analysis is the extraction of data from a laboratory data base with subsequent determination of reference intervals. This is a widely accepted method for determining reference intervals [13,14]. It can be referred to as data mining. It is particularly useful when expensive or difficult testing done in children is to be analysed. Another challenge in determining reference intervals is the accommodation of changes in the population distribution of results due to gender or age. Clearly, if differences in assay results occur in people of different age or gender, and these are not recognised, misinterpretation of results can occur. The LMS analytical method developed by TJ Cole and used to describe growth curves for children effectively allows evaluation of changes in data by age [15].

## 2. Methods

We received 7851 requests for 17 assays of 15 enzymes in leukocyte and plasma lysosomal enzyme assays were identified from our laboratory databases between January 1985 and September 2013. Results were excluded if they were repeat/duplicate samples, known to be affected (i.e. monitoring samples), or were known to be pseudo-deficient. Data was accessed in the study under and institutional review board waiver for use of data under quality assurance purposes, in this case specifically for the development of reference intervals. The institutional review board is that of South Australia (SA) Pathology.

### 2.1. Enzyme assays

Assays for the lysosomal enzyme-screening panel are as follows, with the associated disease being denoted in table 1. Methods are also referenced in the table, where published. Total beta-hexosaminidase [16], palmitoyl protein thioesterase 1 [17], beta-hexosaminidase A [18], beta-glucuronidase [19], beta-galactosidase [19], acid phosphatase [19], alpha-mannosidase [19], alpha-fucosidase [19], beta-glucuronidase [19] and alpha-N-acetylgalactosaminidase [16] are performed using artificial 4-methylumbelliferone (4MU) conjugated substrates.

**Table 1.** Reference intervals for the lysosomal enzyme-screening panel in unaffected individuals.

| Enzyme | Disease (MIM #) | Specimen | Method | N | /min/mg<br>protein or<br>*/min/m<br>L | Old<br>LRL | New<br>LRL | New<br>LRL<br>CI | Old<br>URL | New<br>URL | New<br>URL<br>CI | Old<br>affecte<br>d<br>range | Ref |
| --- | --- | --- | --- | --- | --- | --- | --- | --- | --- | --- | --- | --- | --- |
| Acid lipase | Cholesteryl ester storage disease; Wolman disease (278000) | L | R | 3955 | pmol | 0.60 | 0.70 | 0.66 to 0.72 | 6.20 | 5.01 | 4.90 to 5.20 | < 0.2 | [20] |
| Acid phosphatase | Lysosomal acid phosphatase deficiency (200950) | L | F | 3066 | nmol | 20.00 | 49.00 | 45 to 53 | 360.00 | 340.00 | 330 to 350 | < 10.0 | [16] |
| Acid Sphingomyelinase | Niemann-Pick disease, type A / B (257200; 607616) | L | R | 6701 | pmol | 20.00 | 16.00 | 15 to 16 | 120.00 | 100.00 | 97 to 100 | < 5 | [26] |
| Alpha-fucosidase | Fucosidosis (230000) | L | F | 6722 | *nmol | 0.40 | 0.58 | 0.56 to 0.60 | 2.70 | 2.70 | 2.70 to 2.80 | <0.05 | [17] |
| Alpha-mannosidase | Mannosidosis, alpha-, types I and II (248500) | L | F | 6734 | *nmol | 2.00 | 4.30 | 4.10 to 4.50 | 25.00 | 23.00 | 22.00 to 23.00 | < 0.05 | [18] |
| Alpha-mannosidase | Mannosidosis, alpha-, types I and II (248500) | P | F | 6000 | *nmol | 0.50 | 0.12 | 0.11 to 0.12 | 3.00 | 2.50 | 2.50 to 2.60 | < 0.03 | [19] |
| Alpha-N-acetylgalactosaminidase | Schindler disease, types I and III (609241); Kanzaki disease (609242) | P | F | 6704 | *nmol | 0.24 | 0.10 | 0.10 to 0.10 | 1.40 | 2.50 | 2.50 to 2.60 | < 400 | U |
| Arylsulphatase A | Metachromatic leukodystrophy (250100) | L | S | 6644 | nmol | 1.00 | 0.90 | 0.80 to 1.00 | 5.00 | 4.90 | 4.90 to 5.10 | < 0.2 | [19] |
| Beta-galactocerebrosidase | Krabbe disease (245200) | L | R | 5796 | pmol | 2.00 | 2.20 | 2.10 to 2.20 | 30.00 | 25.00 | 24.00 to 26.00 | < 0.5 | U |
| Beta-galactosidase | GM1-gangliosidosis, type I (230500) | L | F | 6733 | nmol | 1.00 | 1.30 | 1.30 to 1.40 | 6.00 | 6.00 | 5.90 to 6.10 | <0.35 | [21] |
| Beta-glucocerebrosidase | Gaucher disease, types I, II and III (230800) | L | R | 5787 | pmol | 600.00 | 617.00 | 607 to 630 | 3200.00 | 2700.00 | 2700 to 2800 | - | [19] |
| Beta-glucuronidase | Mucopolysaccharidosis VII (253220) | L | F | 6723 | nmol | 0.50 | 0.53 | 0.52 to 0.55 | 3.40 | 3.10 | 3.10 to 3.20 | <0.03 | U |
| Beta-glucuronidase | Mucopolysaccharidosis VII (253220) | P | F | 755 | *nmol | 0.10 | 0.08 | 0.07 to 0.09 | 2.00 | 1.10 | 0.93 to 1.40 | < 0.5 | [19] |
| Beta-hexosaminidase A | GM2-gangliosidosis, Tay-Sachs disease (272800) | L | F | 6723 | nmol | 2.00 | 4.00 | 3.80 to 4.10 | 20.00 | 15.00 | 15.00 to 15.00 | < 0.1 | [19] |
| Palmitoyl protein thioesterase 1 | Neuronal ceroid lipofuscinosis, type 1 (256730) | L | F | 2476 | nmol | 0.50 | 0.54 | 0.50 to 0.60 | 2.00 | 2.30 | 2.20 to 2.50 | < 0.1; > 7.0 | [19] |
| Total beta-hexosaminidase | Sandhoff disease (268800) | L | F | 6732 | nmol | 21.50 | 15.00 | 14 to 15 | 51.00 | 49.00 | 48 to 50 | - | [16] |
| Tripeptidyl | Neuronal ceroid | L | F | 2471 | nmol | 0.80 | 0.66 | 0.60 | 2.00 | 1.90 | 1.80 t | > 25 | [19] |
| Peptidase 1 | lipofuscinosis, type<br>2 (204500) |  |  |  |  |  |  | to<br>0.70 |  |  | o 1.90 |  |  |

L, leukocytes; P, plasma; F, fluorometric; R, radionucleotide; S, spectrophotometric; U, unpublished in-house method, described in methods.

Tripeptidyl peptidase 1 is assayed using a fluorogenic 7-amino-4-methylcoumarin conjugated substrate [20]. Arylsulphatase A activity is determined spectrophotometrically using a synthetic p-nitrocatechol sulphate substrate [21]. The remainder of the enzymes are assayed using radiolabelled substrates that are synthesized in-house Acid lipase [32] and acid sphingomyelinase [33] activity were determined using published methods as described. The beta-galactocerebrosidase and beta-glucocerebroside methods are described below.

#### 2.1.1. Beta-galactocerebrosidase

The beta-galactocerebrosidase substrate, [^3^H]galactosylceramide, is synthesised as described in (Y. Suzuki *et al*, <u>J. Lip. Res</u>. (1972) **13**: 687-690). The working substrate stock was made by the addition of 1.0 mL [^3^H]galactosylceramide (approximately 7000 dpm/uL), 0.05 mL 0.1μmol/mL bovine brain cerebrosides, (Sigma; in chloroform:methanol (2:1, v/v)). 1.0 mL 1 mg/mL oleic acid, (Unilab in chloroform:methanol (2:1, v/v)) and 1.0 mL 70 mg/mL pure sodium taurocholate (Calbiochem; in chloroform:methanol (2:1, v/v)). Each reaction contained 31uL [^3^H]galactosylceramide working substrate stock dried down under nitrogen stream in the reaction tube. 25uL 0.2 mol/L sodium acetate buffer, pH 4.6 and 75μL leucocyte extract (60-80 μg protein) were then added. After a 3 hour incubation period at 37°C, 2.0 ml chloroform methanol (2:1, v/v), 0.4mL water and 10μL 1mg/mL galactose was added. The mixtures were vortexed and centrifuged for 5 min at 500 x g. Released [3H]galactose was measured in 500 uL aliquots of the upper phase by mixing with 4 mL Optiphase ‘Hisafe’ 3 (Wallac) scintillation fluid and quantified by liquid scintillation counting.

#### 2.1.2. Beta-glucocerebrosidase

##### 2.1.2.1 Substrate synthesis

[^14^C]-ceramide-beta-glucoside was synthesized in-house and prepared as follows. 40 mg anhydrous glucopsychosine (lyso-glucocerebroside; Matreya) was refluxed at 105o for 16 h with 2.1 mL 5 N (aqueous) potassium hydroxide and 19 mL n-butanol. 22 mL of water and 42 mL of chloroform-methanol (2:1, v/v) were added after the reaction mixture cooled to room. The lower phase was evaporated to dryness at 40o, then the residue dissolved in a small volume of chloroform-methanol (2:1, v/v) and applied to a silica gel 60 plate (Merck). The chromatogram was resolved in chloroform-methanol-water (65:25:3, v/v/v). The lysoglucocerebroside zone was visualised by exposure to iodine vapour, scraped from the plate and eluted with 30 mL chloroform-methanol-acetic acid-water (70:30:1:3, v/v/v/v). This was evaporated to dryness under nitrogen at 40o, then dissolved in a small volume of chloroform-methanol (2:1, v/v) and centrifuged. The supernatant was transferred to a new tube. This procedure was repeated until the dissolved silica was removed. 250 μCi [1-14C] stearic acid (59 mCi/mmol, New England Nuclear) was evaporated to dryness three times in the presence of benzene previously dried over sodium metal. 0.25 mL dry benzene and 0.25 ml oxalyl chloride (Sigma) were added to the tube containing [1-14C] stearic acid and allowed to stand at 37o for 1 h. This was then dried under nitrogen at 40o and a solution of lysoglucocerebroside (4.5 umol) in 600 μL mixture of freshly distilled tetrahydrofuran over lithium aluminium hydride (Sigma) and 150 mM aqueous sodium carbonate (22:10, v/v) was added. This was mixed and allowed to stand at room temperature for 50 min, then dried under nitrogen at 40oC. 3 mL of chloroform-methanol (2:1, v/v) and 0.75 ml of water was added to the residue and vortexed. The lower phase was evaporated to dryness and the residue was partitioned again in the same way. The residue, containing [^14^C]-ceramide-beta-glucoside, was dissolved in a small volume of chloroform-methanol (2:1 v/v) and applied to a silica gel 60 plate. After chromatography in chloroform-methanol-water (90:10:1, v/v), the [^14^C]-ceramide-beta-glucoside was located by radioautography and eluted from the silica with 5 ml chloroform-methanol-water (60:30:5, v/v/v). To prepare the working substrate stock, 0.5mL [^14^C]-ceramide-beta-glucoside (approximately 3500-4000 dpm/uL), 0.5mL 5mg/mL Triton X-100 (BDH; prepared in cholorform-methanol (2:1, v/v)), 0.125mL 100mg/mL sodium taurocholate (BDH; dissolved in cholorform-methanol (2:1, v/v)) and 0.1mL 0.4mg/mL bovine glucocerebroside (Matreya; dissolved in dissolved in cholorform-methanol (2:1, v/v)) were mixed together.

##### 2.1.2.2 Beta-glucocerebrosidase assay

Each reaction contained 35uL [^14^C]-ceramide-beta-glucoside working substrate stock, dried down under nitrogen stream, resuspended in 50uL water and sonicated briefly to dissolve. To this, 10μL citrate-phosphate buffer, pH 5.2 and 50μL leucocyte extract (20-40 μg protein) were added and incubated for 3 hours at 37°C. 500 μL chloroform:methanol (1:1, v/v) and 125μL water were added and the mixtures vortexed and centrifuged for 5 min at 500 x g. The upper phase was removed and the lower phase was dried down under nitrogen, then the residue resuspended in 10μL chloroform:methanol (2:1, v/v). 5μL of each reaction, together with 1uL of chromatography markers (5mg/mL each of ceramide (Matreya) and glucocerebroside (Matreya) in chloroform:methanol (2:1, v/v)) were applied to a silica gel 60 plate. Following chromatography in chloroform:methanol:water (45:5:0.5, v/v/v/v), the plate was dried and exposed to iodine vapour to expose the markers. Each reaction lane was cut into 2 pieces to separate the marker bands and transferred to separate vials with 1mL methanol to elute. 4 ml Optiphase ‘Hisafe’ 3 (Wallac) scintillation fluid was added and released [^14^C]ceramide was quantified by liquid scintillation counting. Beta-glucocerebrosidase activity is determined by measuring substrate to product conversion.

### 2.2. Calculation of reference intervals

Reference intervals were calculated using MedCalc for Windows version 12.5 (MedCalc Software, Ostend, Belgium), according to the Clinical and Laboratory Standards Institute C-28A protocol (CSLI-C28A) [27], using the non-parametric percentile method, with 95% reference intervals and 95% confidence intervals (CI) of the upper and lower reference limits (URL and LRL). Age and sex partitioning were explored using LMS software the LMS ChartMaker Light software, version 2.3.

### 2.3. Affected values

Values for affected patients were extracted from the institutional database and descriptive statistics are included.

## 3. Results

New and old reference intervals for the seventeen assays are displayed in table 1. Reference values for our cohort of affected individuals are displayed in table 2. Reference intervals were determined by analyzing in excess of 7500 results for most assays. These were collected over a period of greater than 15 years and across a wide distribution of age in both genders. More than 75% of the assays were performed in children less than 16 years of age.

**Table 2.** Reference intervals for the lysosomal enzyme-screening panel in affected individuals.

| Enzyme | Disease (MIM #) | Specimen | Activity Units /min/mg protein or */min/ml | N | Minimum value | Median | Maximum value |
| --- | --- | --- | --- | --- | --- | --- | --- |
| Acid lipase | Cholesteryl ester storage disease; Wolman disease (278000) | L | pmol | 19 | 0 | 0.03 | 0.23 |
| Acid Sphingomyelinase | Niemann-Pick disease, type A/B (257200; 607616) | L | pmol | 22 | 0.17 | 2.25 | 7.6 |
| Alpha-fucosidase | Fucosidosis (230000) | L | *nmol | 3 | 0 | 0 | 0.04 |
| Alpha-mannosidase | Mannosidosis, alpha-, types I and II (248500) | L | *nmol | 13 | 0.09 | 0.14 | 0.47 |
| Alpha-mannosidase | Mannosidosis, alpha-, types I and II (248500) | P | *nmol | 43 | 15 | 55 | 120 |
| Alpha-N-acetylgalactosaminidase | Schindler disease, types I and III (609241); Kanzaki disease (609242) | P | *nmol | 1 | 0.1 | 0.1 | 0.1 |
| Arylsulphatase A | Metachromatic leukodystrophy (250100) | L | nmol | 61 | 0 | 0.13 | 0.57 |
| Beta-galactocerebrosidase | Krabbe disease (245200) | L | pmol | 27 | 0 | 0.12 | 0.4 |
| Beta-galactosidase | GM1-gangliosidosis, type I (230500) | L | nmol | 33 | 0.01 | 0.09 | 0.22 |
| Beta-glucocerebrosidase | Gaucher disease, types I, II and III (230800) | L | pmol | 99 | 0 | 140 | 425 |
| Beta-glucuronidase | Mucopolysaccharidosis VII (253220) | L | nmol | 3 | 0.01 | 0.03 | 0.04 |
| Beta-glucuronidase | Mucopolysaccharidosis VII (253220) | P | *nmol | 43 | 3.6 | 20 | 46 |
| Beta-hexosaminidase A | GM2-gangliosidosis; Tay-Sachs disease (272800) | L | nmol | 31 | 0.05 | 0.14 | 0.81 |
| Palmitoyl protein thioesterase 1 | Ceroid lipofuscinosis, neuronal, 1 (256730) | L | nmol | 7 | 0.1 | 0.3 | 0.4 |
| Total beta-hexosaminidase | Sandhoff disease (268800) | L | nmol | 20 | 0.78 | 1.85 | 2.9 |
| Tripeptidyl Peptidase 1 | Ceroid lipofuscinosis, neuronal, 2 (204500) | L | nmol | 42 | 0 | 0.02 | 0.08 |

With the exception of the alpha-fucosidase URL, arylsulphatase A LRL and palmitoyl protein thioesterase 1 LRL, the newly calculated reference intervals varied significantly from the historic values, i.e. the historic value fell outside the 95% CI for the newly calculated reference limit. Despite this, a substantial difference remained between the new reference limits and the levels observed in affected individuals. No useful differences were noted with age or gender partitions.

## 4. Discussion

### 4.1. Nature of patient reference sample

This paper reports reference intervals for enzyme activity in 17 assays of 15 enzymes used for the evaluation of LSDs. These reference intervals have been determined by the analysis of more than 7500 results for most assays. These results span a period of greater than 15 years and across a wide distribution of age in both genders. More than 75% of the assays were performed in children less than 16 years of age.

These assays were performed to investigate patients with clinical features prompting consideration of a diagnosis of a lysosomal storage disorder. However, as each enzyme is tested as part of a panel, the overwhelming majority of tests were performed in people with little likelihood of the specific lysosomal storage disorder tested by that assay. It may even be advantageous that these reference intervals are generated from patient data, as it will allow differentiation of affected patients from those for whom an LSD may be suspected [28,29].

The results of patients known to be affected were excluded.

### 4.2. Calculation of reference intervals

The development of reference intervals is critical for the interpretation of laboratory tests, especially in uncommon and poorly harmonized tests, such as the measurement of enzyme activity associated with neuronopathic LSDs. Reference intervals are particularly difficult to determine for small volume, expensive and laborious assays that are used in children. The preferred method for determining reference intervals is in an *a priori* or direct fashion using a reference population of suitable age and gender distribution known to be in good health. This is difficult and methods for developing reference intervals in an *a posteriori* or indirect fashion are increasingly used [28,30,31].

The reference intervals described are the 95 % confidence intervals of the central 95 % distribution. While one might use a central 99 % distribution, the 95 % allows a more standard understanding of results and militates against false negative reports [27]. The central 95 % is recommended by CLSI [27].

### 4.3. Partitioning of reference intervals by age and sex

Two x-linked LSDs are known, neither of which are included in this panel of enzyme assays. There has not been any sex-bias reported with respect to clinical presentation, metabolite levels or enzyme activities in any of the other LSDs. Age-bias is apparent in the sense that childhood is the most common period for these disorders to present, and adult presentations remain relatively rare. No clinically relevant variations were observed in our unaffected cohort after the data sets for each assay were partitioned by gender. Likewise, assessment of the results distributed by age did not reveal any clinically relevant variations. Therefore use of a single reference interval for each white cell assay is appropriate.

### 4.4. Benefits of acquisition of reference intervals

The CLSI guidelines for determining reference intervals from a reference population were used to determine reference intervals for the screening panel. While these reference intervals are not based on patient outcome studies, the highest standard for reference intervals based on the Stockholm criteria, they are based on careful analysis of a reference population of unaffected individuals, the second highest grade. These reference intervals replace reference intervals whose origin is simply unknown. The newly developed reference intervals can be applied confidently to the lysosomal enzyme screening panel and be used to evaluate panel results informing further investigation of the patients studied. There is no longer a concern that the results are compared to some historical, unproven reference intervals. Additionally, ISO 15189 states that “the laboratory shall define biological reference intervals or clinical decision values, document the basis for the reference intervals or decision limits and communicate this information to users”. These assays are now ISO 15189 compliant as the source of the reference intervals is known, documented, and communicated.

## 5.0 Conclusion

We have developed reference intervals for a suite of lysosomal enzyme assays based on a posteriori, non-parametric analysis following CLSI guidelines using patient data including results from men, women, and children from the laboratory database. The analysis shows that single reference intervals can be used for all patients for each assay. Having developed evidence based reference intervals using the Stockholm Criteria, we can now confidently report assays that are more ISO 15189 compliant.

## Data Availability

Not available.

## Acknowledgments

The authors wish to thank Enzo Ranieri for useful discussions and Elizabeth Gjerde for facilitating the quality assurance process at SA Pathology.

## Author Contributions

MM and JF conceived and supervised the project. DM and SS performed the data extraction. DM and MM performed the statistical analysis and wrote the manuscript drafts. SS and MF wrote the laboratory methods section. DM, MM and SS were involved in writing and editing of the manuscript.

## Conflicts of Interest

The authors declare no conflict of interest.

## Notes

### Competing Interest Statement

The authors have declared no competing interest.

### Funding Statement

Work funded through employment with SA PAthology, a public laboratory.

### Author Declarations

Data was accessed in the study under and institutional review board waiver for use of data under quality assurance purposes, in this case specifically for the development of reference intervals. The institutional review board is that of South Australia (SA) Pathology.

## References

1. Brady, R. The concept of treatment in lysosomal storage diseases. In Lysosomal storage disorders, Springer US: 2007; pp 37–43.

2. Generoso, A.; Giuseppe, L. Diagnosis and management of lysosomal storage disorders. Three key words: Early, multidisciplinary, and network. Cardiogenetics 2013, 3, e1–e1.

3. Platt, F.; Butters, T. Substrate reduction therapy. In Lysosomal storage disorders, Springer US: 2007; pp 153–168.

4. Cox, T.M. Current treatments. In Lysosomal storage disorders, John Wiley & Sons, Ltd: 2012; pp 151–165.

5. Valayannopoulos, V.; Brassier, A.; Chabli, A.; Caillaud, C.; Lemoine, M.; Odent, T.; Arnoux, J.B.; de Lonlay, P. [enzyme replacement therapy for lysosomal storage disorders]. Arch Pediatr 2011, 18, 1119–1123.

6. Andrew Burrow, T.; Grabowski, G.A. Emerging treatments and future outcomes. In Lysosomal storage disorders, John Wiley & Sons, Ltd: 2012; pp 174–180.

7. Garber, K. Orphazyme: This danish startup intends to use a chaperone protein to treat lysosomal storage disorders. Nature Biotechnology 2013, 31, 189.

8. Edmond Wraith, J.; Beck, M. Clinical aspects and clinical diagnosis. In Lysosomal storage disorders, John Wiley & Sons, Ltd: 2012; pp 13–19.

9. Jardim, L.B.; Villanueva, M.M.; Souza, C.F.M.d.; Netto, C.B.O. Clinical aspects of neuropathic lysosomal storage disorders. Journal of Inherited Metabolic Disease 2010, 33, 315–329.

10. Wood, T.; Basehore, M.; Jones, J.; Friez, M.; Cathey, S.; Pollard, L. Development of a next generation sequencing panel for lysosomal storage disorders. Molecular genetics and metabolism 2013, 108, S100.

11. Fraser, C.G.; Kallner, A.; Kenny, D.; Petersen, P.H. Introduction: Strategies to set global quality specifications in laboratory medicine. Scandinavian journal of clinical and laboratory investigation 1999, 59, 477–478.

12. Sikaris, K. Application of the stockholm hierarchy to defining the quality of reference intervals and clinical decision limits. The Clinical biochemist. Reviews / Australian Association of Clinical Biochemists 2012, 33, 141–148.

13. Bock, B.J.; Dolan, C.T.; Miller, G.C.; Fitter, W.F.; Hartsell, B.D.; Crowson, A.N.; Sheehan, W.W.; Williams, J.D. The data warehouse as a foundation for population-based reference intervals. American journal of clinical pathology 2003, 120, 662–670.

14. Shine, B. Use of routine clinical laboratory data to define reference intervals. Annals of clinical biochemistry 2008, 45, 467–475.

15. Cole, T.J.; Green, P.J. Smoothing reference centile curves: The lms method and penalized likelihood. Statistics in medicine 1992, 11, 1305–1319.

16. Leaback, D.H.; Walker, P.G. Studies on glucosaminidase. 4. The fluorimetric assay of n-acetyl-beta-glucosaminidase. The Biochemical journal 1961, 78, 151–156.

17. Baum, H.; Dodgson, K.S.; Spencer, B. The assay of arylsulphatases a and b in human urine. Clinica chimica acta; international journal of clinical chemistry 1959, 4, 453–455.

18. Inui, K.; Wenger, D.A. Usefulness of 4-methylumbelliferyl-6-sulfo-2-acetamido-2-deoxy-beta-d-glucopyrano sid e for the diagnosis of gm2 gangliosidoses in leukocytes. Clinical genetics 1984, 26, 318–321.

19. Kolodny, E.H.; Mumford, R.A. Human leukocyte acid hydrolases: Characterization of eleven lysosomal enzymes and study of reaction conditions for their automated analysis. Clinica chimica acta; international journal of clinical chemistry 1976, 70, 247–257.

20. Sohar, I.; Sleat, D.E.; Jadot, M.; Lobel, P. Biochemical characterization of a lysosomal protease deficient in classical late infantile neuronal ceroid lipofuscinosis (lincl) and development of an enzyme-based assay for diagnosis and exclusion of lincl in human specimens and animal models. Journal of neurochemistry 1999, 73, 700–711.

21. Hultberg, B. Fluorometric assay of the arylsulphatases in human urine. Journal of clinical chemistry and clinical biochemistry. Zeitschrift fur klinische Chemie und klinische Biochemie 1979, 17, 795–797.

22. Dubois, G.; Zalc, B.; Le Saux, F.; Baumann, N. Stearoyl[1-14c]sulfogalactosylsphingosine ([14c]sulfatide) as substrate for cerebroside sulfatase assay. Analytical biochemistry 1980, 102, 313–317.

23. Kudoh, T.; Wenger, D.A. Diagnosis of metachromatic leukodystrophy, krabbe disease, and farber disease after uptake of fatty acid-labeled cerebroside sulfate into cultured skin fibroblasts. The Journal of clinical investigation 1982, 70, 89–97.

24. Poulos, A.; Beckman, K. Trihexosylceramide alpha-galactosidase of human leucocytes. Clinica chimica acta; international journal of clinical chemistry 1978, 89, 35–45.

25. Suzuki, Y.; Suzuki, K. Specific radioactive labeling of terminal n-acetylgalactosamine of glycosphingolipids by the galactose oxidase-sodium borohydride method. Journal of lipid research 1972, 13, 687–690.

26. Poulos, A.; Beckman, K. The bile salt activation of leucocyte sphingolipid hydrolase activity and the modifying effects of triton x-100. Clinica chimica acta; international journal of clinical chemistry 1980, 107, 27–35.

27. Horowitz, G.L.; Clinical and Laboratory Standards Institute. Defining, establishing, and verifying reference intervals in the clinical laboratory : Approved guideline. 3rd ed.; Clinical and Laboratory Standards Institute: Wayne, Pa., 2008; p 61 p.

28. Katayev, A.; Balciza, C.; Seccombe, D.W. Establishing reference intervals for clinical laboratory test results: Is there a better way? American journal of clinical pathology 2010, 133, 180–186.

29. Kouri, T.; Kairisto, V.; Virtanen, A.; Uusipaikka, E.; Rajamaki, A.; Finneman, H.; Juva, K.; Koivula, T.; Nanto, V. Reference intervals developed from data for hospitalized patients: Computerized method based on combination of laboratory and diagnostic data. Clinical chemistry 1994, 40, 2209–2215.

30. Concordet, D.; Geffre, A.; Braun, J.P.; Trumel, C. A new approach for the determination of reference intervals from hospital-based data. Clinica chimica acta; international journal of clinical chemistry 2009, 405, 43–48.

31. Koerbin, G.; Sikaris, K.A.; Jones, G.R.; Ryan, J.; Reed, M.; Tate, J.; On behalf of the, A.C.f.C.R.I. Evidence-based approach to harmonised reference intervals. Clinica chimica acta; international journal of clinical chemistry 2013.

32. Burton BK, Emery D Mueller HW. Lysosomal acid lipase in cultivated fibroblasts: characterisation of enzyme activity in normal and enzymatically deficient cell lines. Clin. Chim. Acta 1980, 101, 25–92.

33. Poulos A, Beckman K. The bile salt activation of leucocyte sphingolipid hydrolase activity and the modifying effects of triton X-100. Clin Chim Acta. 1980 107, 1–2, 27-35.

